# Identifying Patient Subpopulations with Significant Race-Sex Differences in Emergency Department Disposition Decisions

**DOI:** 10.1101/2024.05.12.24307254

**Authors:** Peter Lin, Nilay T Argon, Qian Cheng, Christopher S Evans, Benjamin Linthicum, Yufeng Liu, Abhishek Mehrotra, Laura Murphy, Mehul D Patel, Serhan Ziya

## Abstract

**Background/Objectives:** The race-sex differences in emergency department (ED) disposition decisions have been reported widely. Our objective is to identify demographic and clinical subgroups for which this difference is most pronounced, which will facilitate future targeted research on potential disparities and interventions.

**Methods:** We performed a retrospective analysis of 93,987 White and African-American adults assigned an Emergency Severity Index of 3 at three large EDs from January 2019 to February 2020. Using random forests, we identified the Elixhauser comorbidity score, age, and insurance status as important variables to divide data into subpopulations. Logistic regression models were then fitted to test race-sex differences within each subpopulation while controlling for other patient characteristics and ED conditions.

**Results:** In each subpopulation, African-American women were less likely to be admitted than White men with odds ratios as low as 0.304 (95% confidence interval (CI): [0.229, 0.404]). African-American men had smaller admission odds compared to White men in subpopulations of 41+ years of age or with very low/high Elixhauser scores, odds ratios being as low as 0.652 (CI: [0.590, 0.747]). White women were less likely to be admitted than White men in subpopulations of 18-40 or 41-64 years of age, with low Elixhauser scores, or with Self-Pay or Medicaid insurance status with odds ratios as low as 0.574 (CI: [0.421, 0.784]).

**Conclusions:** While differences in likelihood of admission were lessened by younger age for African-American men, and by older age, higher Elixhauser score, and Medicare or Commercial insurance for White women, they persisted in all subgroups for African-American women. In general, patients of age 64 years or younger, with low comorbidity scores, or with Medicaid or no insurance appeared most prone to potential disparities in admissions.

## INTRODUCTION

The disposition decision is an important timepoint during a patient’s visit to an emergency department (ED) as it represents a split in the patient’s path through the healthcare system. Even though patients may no longer remain within an ED after receiving a disposition, this decision continues to affect their health in the long term^1–3^. Consequently, patients facing disparities in disposition decisions may be impacted long after they depart the ED, and hence, it is of interest to study differences and disparities surrounding disposition decisions closely. An American College of Emergency Physicians workgroup convened in 2021 identified the measurement of disparities in ED processes, including disposition decision, as a priority to promote health equity in emergency medicine^4^.

Disparities have been shown to affect ED patients in many different decisions and specific situations^5–7^. Opioid prescribing practices have been documented to vary by patient race^8–10^. Patients, both adult and pediatric, suffering from chest pain, have received different treatments depending on sex, race, and insurance status^11–12^. Those who display potential acute coronary syndrome received different care depending on their sex^13^ or if they were White or African American^14–15^. In addition, prior work reports demographic differences in triage score assignment^16–19^ as well as rooming prioritization^20–21^, with African-American and Hispanic patients receiving less acute scores and being less likely to be prioritized for rooming.

Several studies have also investigated differences and disparities surrounding ED disposition decisions. In particular, women, African Americans, and uninsured patients have been found to be less likely to be admitted from the ED compared to men, Whites, and privately insured patients^22–28^, respectively. Furthermore, uninsured and Medicaid patients and women are more likely to leave an ED against medical advice^29–30^. To date, much of the literature on differences in ED disposition has focused on specific conditions or complaints, such as chest pain and injury, or adjusting by broad groupings of clinical factors. There is limited evidence on patient subgroups prone to differing ED disposition decisions, which would be invaluable to understanding the causes of these differences and to tailoring and targeting interventions to address them. Given ED patients’ demographic and clinical profiles are highly heterogeneous, machine learning methods, such as random forests, can improve the performance of predictive models^31^. In this retrospective study of adult, intermediate-acuity patients presenting to the ED, we examined the extent of race and sex differences in ED disposition decisions within important demographic and clinical subgroups identified by random forests. Specifically, we tested the null hypothesis that the disposition decision (admit or discharge) did not vary by patient race-sex combination within important patient subgroups when controlling for patient-level and ED operational characteristics. Our objective was to identify subgroups in which race-sex differences were most pronounced to facilitate future research that evaluates potential disparities in admission decisions and explores tailored interventions.

## METHODS

We performed a retrospective analysis of data from three EDs in the Southeastern United States from January 1, 2019 through February 29, 2020. ED A is part of a large academic medical center, containing 59 beds divided among five adult care areas and one pediatric care area. ED B is part of a large community hospital in an urban area, with 48 beds split among four care areas. ED C is part of a large hospital serving a predominantly African-American population in both urban and rural areas. It has 38 beds split between a main care area, a fast-track area for low acuity patients and separate dedicated areas for behavioral health, high intensity, and trauma patients. Notably, the county where ED C is located had a median household income of $55,956 in 2021 compared to respective median household incomes of $79,814 and $91,558 for the counties where ED A and B are located^32^.

Patients were included in the analysis if they were adults (at least 18 years old) who were assigned an Emergency Severity Index (ESI) score of 3 (intermediate acuity). These patients have a similar level of severity and hence are expected to receive similar treatment from decision makers. In addition, the majority of patients both within the study EDs and in general^33^ receive an ESI 3 score at triage. Additional eligibility criteria included being seen in one of the main ED care areas, having complete event timestamps in the electronic health record data, being White or African American, and being admitted to the hospital or discharged from the ED. Interfacility transfers were excluded. Less than 0.01% of patient encounters had a documented sex other than male or female, and these were excluded due to the low patient counts within those demographics affecting model parameter estimates. While our data included three data elements related to sex: “sex,” “gender identity,” and “sex assigned at birth,” we used “sex” since the other two data elements had several missing entries (around 40% of all patient encounters).

### Primary outcome and covariates

The primary outcome in all our models was whether a given patient was admitted versus discharged at the end of an ED visit. Differences in disposition decisions were evaluated between four race-sex groups: African-American female, African-American male, White female, and White male (referent).

In all our models, we adjusted for covariates that were deemed to correlate with admission decisions: age, ethnicity, chief complaint, insurance status, weighted Elixhauser score, care area where disposition was decided, day of week, hour of day, and average waiting time in the ED (as a proxy for crowding level). Patient age was categorized into three groups: 18-40, 41-64, 65 and older. The chief complaint for each patient was first grouped into one of 17 categories^34^, which we further reduced to 13 by combining “General/Minor” and “Environmental” as well as merging the three different “ENT” categories. Comorbidity status (in the form of a comorbidity score, existence or number of chronic conditions, etc.) was found to be a significant independent variable in prior work on demographic differences in ED decisions^12,16,19–20^. In this study, we used weighted Elixhauser comorbidity score^35–36^ as a measure of a patient’s comorbidity burden and case complexity due to its statistical efficiency and relevance demonstrated by our earlier work^19–20^. We calculated the Elixhauser score for each patient encounter by running the R comorbidity package^37^ on the provided ICD-10 codes. We also included insurance status as a covariate to serve as a proxy for level of access to health care. Although insurance status has also been used as a measure of socioeconomic status in prior emergency medicine research, we refrain from making this connection as there is growing evidence indicating that it is does not correlate well with other socioeconomic factors such as income and employment status^38^. For statistical efficiency, we collapsed all patient-level insurance information to reduce the number of required parameters in the models. Specifically, primary and secondary insurance information was used to group patients into a single insurance status variable as follows: patients with “Medicaid” listed for either primary or as the sole secondary insurance were grouped into a “Medicaid” group; Self-Pay patients were grouped together in a separate group. All remaining patients were then placed into “Commercial/Other” or “Medicare,” depending on what their primary insurance was listed as.

We also included several covariates related to ED conditions in addition to patient-specific covariates discussed above. In particular, for each encounter, we noted the hourly block during which the patient received their disposition decision. We then calculated the mean length of time from the end-of-triage to first-seen-by-provider for all patients who entered the waiting room during that same hourly block and used this mean as the ED crowding measure at the time of the disposition decision. Finally, the time of day for disposition decision was recorded as one of six four-hour blocks (12 am to 4 am, 4 am to 8 am, 8 am to 12 pm, 12 pm to 4 pm, 4 pm to 8 pm, and 8 pm to 12 am).

### Statistical analysis

First, using the entire dataset, we fitted a logistic regression model for each ED with all covariates. (Table S1 in the Supplement provides reassuring results of a test that we used to check for potential multicollinearity issues.) In each of these logistic regression models we then computed odds ratios (OR) and 95% confidence intervals (CI) and tested the significance of patient race-sex to determine whether there was evidence of difference, considering *p*-values less than 0.05 to be statistically significant.

To select patient subgroups in which to test race-sex differences, we first fitted a random forest^39–40^ for each ED to obtain variable importance plots that reflect the relative importance of each predictor to model accuracy. (For each random forest, we randomly selected 70% of the corresponding ED’s data for training and used the remaining 30% for testing.) Each random forest comprised of 500 decision trees, each of which was fitted using 4 predictors selected randomly from the entire pool of 10 covariates. The importance of each predictor was determined through the mean decrease in accuracy when that particular variable was permuted in the out-of-bag data. The random forests were fitted using the randomForest^41^ package in R^42^.

Using the variable importance plots, we then selected three of the most important variables for subgroupings: the weighted Elixhauser score, age group, and insurance status. Although the chief complaint category was found to be important consistently across EDs, we did not use it to split our data because of its large number of categories potentially resulting in small sample sizes. Since the weighted Elixhauser score is a continuous variable, groups were set to be 0 and below, 1 through 6, and 7 and above. Age and insurance status groups were set as defined above. For each level we then fitted a logistic regression model using the data belonging to that level with all the covariates except for the one that was being examined. In each of these logistic regression models we then computed ORs and 95% CIs and tested the significance of patient race-sex within each patient subpopulation. All calculations and analysis were performed using R 3.5.3.

This study was reviewed by the IRB at the University of North Carolina at Chapel Hill and determined to be exempt from IRB review (UNC IRB # 22-1714). Upon this IRB exemption, a study dataset was provided to the authors on November 18^th^, 2022. The authors did not have access to information that could identify individual participants. As a retrospective study of existing data with limited identifiers (i.e., patient age and encounter date-times), informed consent was not required.

## RESULTS

Table 1 shows a breakdown of study patient characteristics as well as admission rates for each group. (Table S2 shows breakdowns of additional characteristics.) In total, there were 28,341, 39,048, and 26,598 study patients from EDs A, B, and C with overall admission rates of 35.5%, 30.3%, and 19.2%, respectively. ED B predominantly served White patients and had the oldest population as well as the highest proportion of patients with commercial insurance at 53.9% compared to 44.6% and 30.4% at EDs A and C. ED C served predominantly African-American patients and had the lowest admission rates among all study EDs. (The difference in admission rates at the three EDs could be attributable to differences in patient population characteristics, utilization of ED resources, and access to healthcare resources such as urgent care centers in the local area.) We also see that ED C had the highest proportions of patients with Medicaid or Self-Pay insurance status.

**Table 1.**
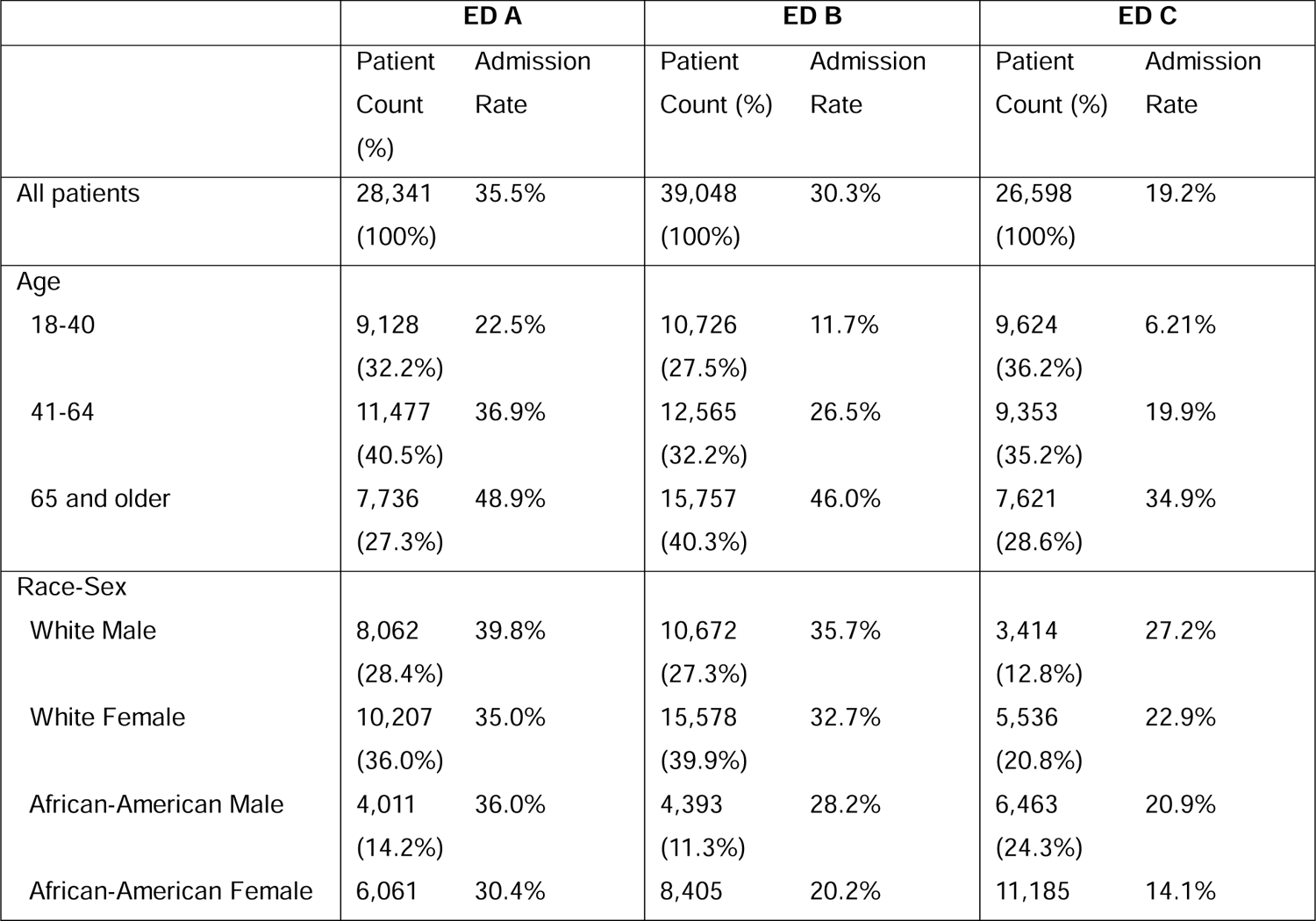
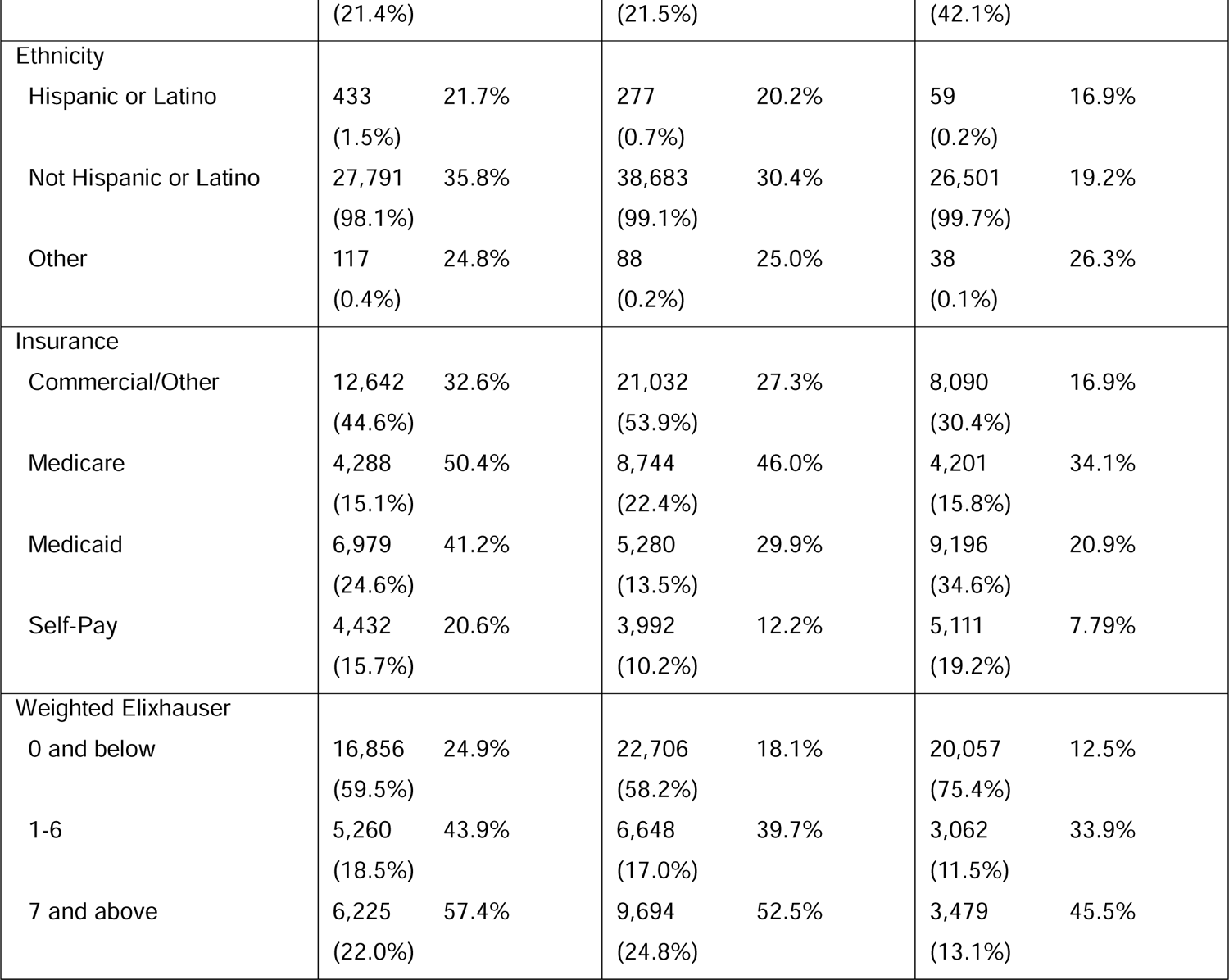
Breakdown of study patient characteristics at each ED with associated admission rates. Admission rates are calculated by dividing the number of patients from each category who were admitted by the total number of patients for that category.

The logistic regression models fitted on the entire datasets for each ED found that all covariates except for patient ethnicity were statistically significant by means of likelihood ratio tests. (The small number of Hispanic patients may explain why ethnicity was the sole insignificant predictor.) The ORs and 95% CIs are provided in Table 2. White men were significantly more likely to be admitted than any other Race-Sex combination at all three EDs: Estimated odds ratios of admission for all other Race-Sex combinations ranged between 0.55 and 0.89 when White men was the reference level, with African-American women having the lowest odds ratios of admission at all three EDs. As expected, the older age groups were more likely to be admitted at EDs A, B, and C with respective odds ratio estimates of 1.68, 2.22, and 2.25 for the 41 through 64 age group and odds ratios of 2.25, 3.88, and 3.25 for the 65 and older group when compared to the 18 through 40 year old group. Of the chief complaint categories, ENT, Genitourinary, Neurologic, Obstetrics/Gynecology, Ophthalmology, Orthopedic, and Trauma patients were significantly more likely to be discharged than Cardiovascular patients at all three EDs. Medicaid and Medicare patients were significantly more likely to be admitted than Commercial/Other patients, with odds ratios ranging from 1.13 to 1.46, and Self-Pay patients were significantly less likely to be admitted with odds ratios ranging between 0.72 and 0.77. Patients with higher Elixhauser scores were more likely to be admitted at all three EDs. At all EDs, the crowding metric was significant, with admission likelihood increasing when the mean waiting time is higher.

**Table 2.**
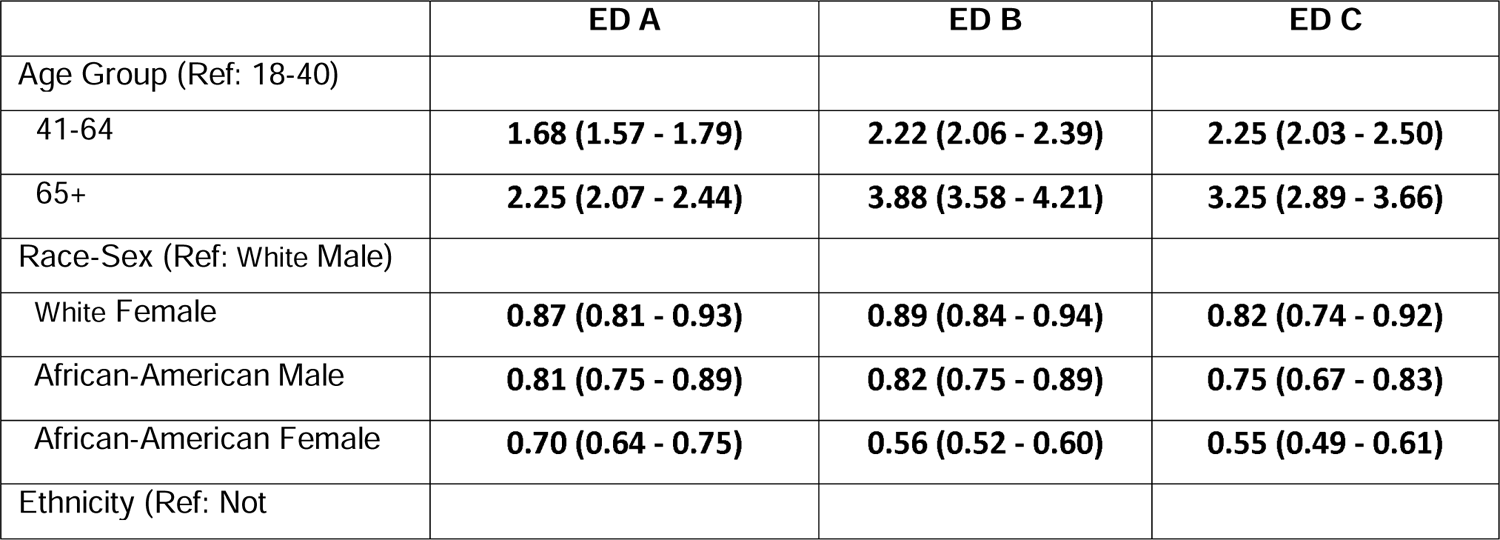
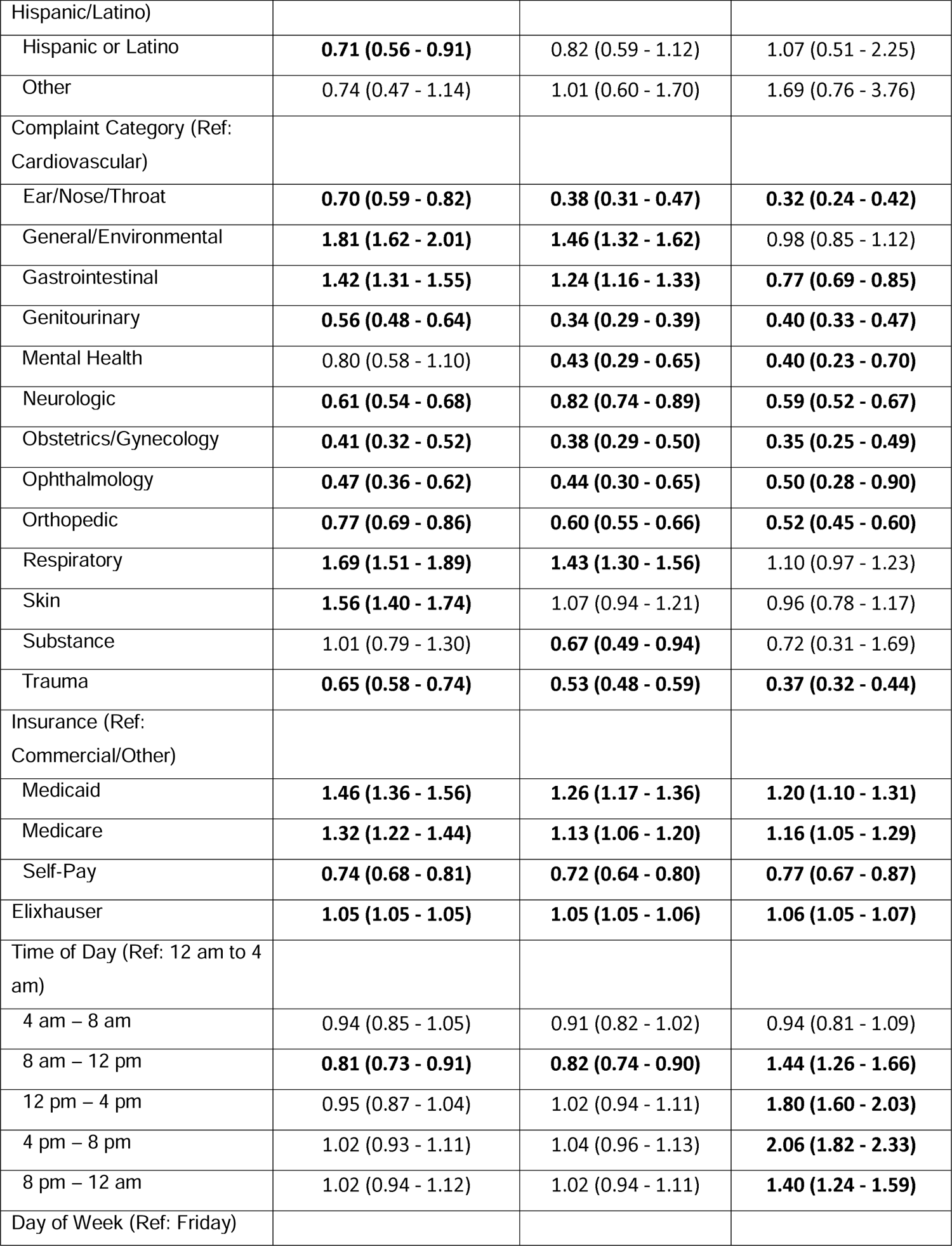
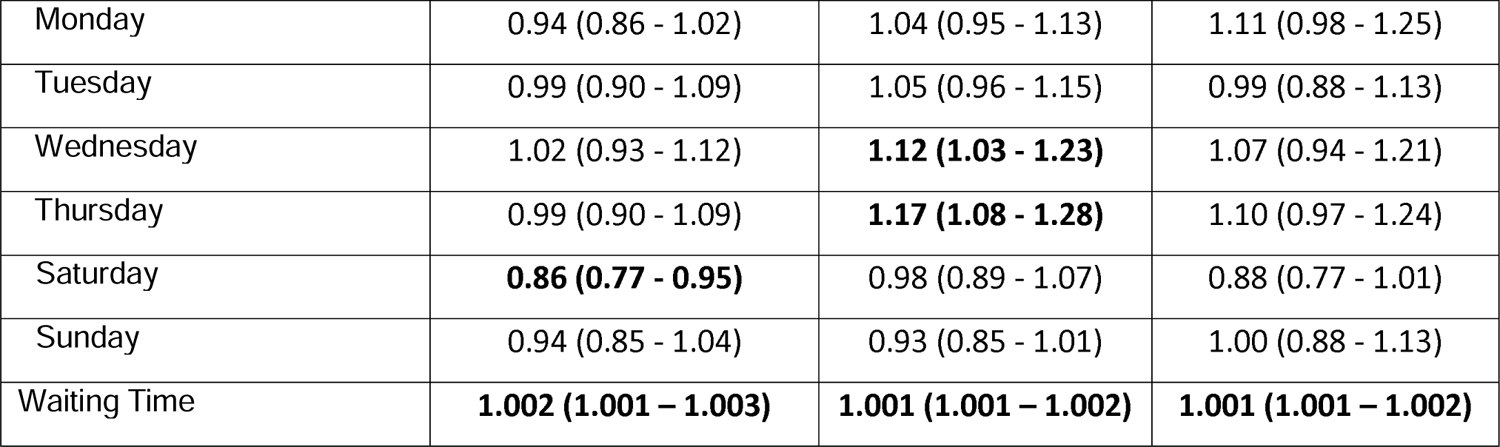
Odds ratios and 95% confidence intervals (CIs) for logistic regression models using all data; statistically significant (p<0.05) CIs are bolded.

|  | **ED A** | **ED B** | **ED C** |
| --- | --- | --- | --- |
| **Age Group (Ref: 18-40)** |  |  |  |
| 41-64 | **1.68 (1.57 - 1.79)** | **2.22 (2.06 - 2.39)** | **2.25 (2.03 - 2.50)** |
| 65+ | **2.25 (2.07 - 2.44)** | **3.88 (3.58 - 4.21)** | **3.25 (2.89 - 3.66)** |
| **Race-Sex (Ref: White Male)** |  |  |  |
| White Female | **0.87 (0.81 - 0.93)** | **0.89 (0.84 - 0.94)** | **0.82 (0.74 - 0.92)** |
| African-American Male | **0.81 (0.75 - 0.89)** | **0.82 (0.75 - 0.89)** | **0.75 (0.67 - 0.83)** |
| African-American Female | **0.70 (0.64 - 0.75)** | **0.56 (0.52 - 0.60)** | **0.55 (0.49 - 0.61)** |
| **Ethnicity (Ref: Not** |  |  |  |
| Hispanic/Latino) |  |  |  |
| Hispanic or Latino | **0.71 (0.56 - 0.91)** | 0.82 (0.59 - 1.12) | 1.07 (0.51 - 2.25) |
| Other | 0.74 (0.47 - 1.14) | 1.01 (0.60 - 1.70) | 1.69 (0.76 - 3.76) |
| Complaint Category (Ref: Cardiovascular) |  |  |  |
| Ear/Nose/Throat | **0.70 (0.59 - 0.82)** | **0.38 (0.31 - 0.47)** | **0.32 (0.24 - 0.42)** |
| General/Environmental | **1.81 (1.62 - 2.01)** | **1.46 (1.32 - 1.62)** | 0.98 (0.85 - 1.12) |
| Gastrointestinal | **1.42 (1.31 - 1.55)** | **1.24 (1.16 - 1.33)** | **0.77 (0.69 - 0.85)** |
| Genitourinary | **0.56 (0.48 - 0.64)** | **0.34 (0.29 - 0.39)** | **0.40 (0.33 - 0.47)** |
| Mental Health | 0.80 (0.58 - 1.10) | **0.43 (0.29 - 0.65)** | **0.40 (0.23 - 0.70)** |
| Neurologic | **0.61 (0.54 - 0.68)** | **0.82 (0.74 - 0.89)** | **0.59 (0.52 - 0.67)** |
| Obstetrics/Gynecology | **0.41 (0.32 - 0.52)** | **0.38 (0.29 - 0.50)** | **0.35 (0.25 - 0.49)** |
| Ophthalmology | **0.47 (0.36 - 0.62)** | **0.44 (0.30 - 0.65)** | **0.50 (0.28 - 0.90)** |
| Orthopedic | **0.77 (0.69 - 0.86)** | **0.60 (0.55 - 0.66)** | **0.52 (0.45 - 0.60)** |
| Respiratory | **1.69 (1.51 - 1.89)** | **1.43 (1.30 - 1.56)** | 1.10 (0.97 - 1.23) |
| Skin | **1.56 (1.40 - 1.74)** | 1.07 (0.94 - 1.21) | 0.96 (0.78 - 1.17) |
| Substance | 1.01 (0.79 - 1.30) | **0.67 (0.49 - 0.94)** | 0.72 (0.31 - 1.69) |
| Trauma | **0.65 (0.58 - 0.74)** | **0.53 (0.48 - 0.59)** | **0.37 (0.32 - 0.44)** |
| Insurance (Ref: Commercial/Other) |  |  |  |
| Medicaid | **1.46 (1.36 - 1.56)** | **1.26 (1.17 - 1.36)** | **1.20 (1.10 - 1.31)** |
| Medicare | **1.32 (1.22 - 1.44)** | **1.13 (1.06 - 1.20)** | **1.16 (1.05 - 1.29)** |
| Self-Pay | **0.74 (0.68 - 0.81)** | **0.72 (0.64 - 0.80)** | **0.77 (0.67 - 0.87)** |
| Elixhauser | **1.05 (1.05 - 1.05)** | **1.05 (1.05 - 1.06)** | **1.06 (1.05 - 1.07)** |
| Time of Day (Ref: 12 am to 4 am) |  |  |  |
| 4 am – 8 am | 0.94 (0.85 - 1.05) | 0.91 (0.82 - 1.02) | 0.94 (0.81 - 1.09) |
| 8 am – 12 pm | **0.81 (0.73 - 0.91)** | **0.82 (0.74 - 0.90)** | **1.44 (1.26 - 1.66)** |
| 12 pm – 4 pm | 0.95 (0.87 - 1.04) | 1.02 (0.94 - 1.11) | **1.80 (1.60 - 2.03)** |
| 4 pm – 8 pm | 1.02 (0.93 - 1.11) | 1.04 (0.96 - 1.13) | **2.06 (1.82 - 2.33)** |
| 8 pm – 12 am | 1.02 (0.94 - 1.12) | 1.02 (0.94 - 1.11) | **1.40 (1.24 - 1.59)** |
| Day of Week (Ref: Friday) |  |  |  |
| Monday | 0.94 (0.86 - 1.02) | 1.04 (0.95 - 1.13) | 1.11 (0.98 - 1.25) |
| Tuesday | 0.99 (0.90 - 1.09) | 1.05 (0.96 - 1.15) | 0.99 (0.88 - 1.13) |
| Wednesday | 1.02 (0.93 - 1.12) | **1.12 (1.03 - 1.23)** | 1.07 (0.94 - 1.21) |
| Thursday | 0.99 (0.90 - 1.09) | **1.17 (1.08 - 1.28)** | 1.10 (0.97 - 1.24) |
| Saturday | **0.86 (0.77 - 0.95)** | 0.98 (0.89 - 1.07) | 0.88 (0.77 - 1.01) |
| Sunday | 0.94 (0.85 - 1.04) | 0.93 (0.85 - 1.01) | 1.00 (0.88 - 1.13) |
| Waiting Time | **1.002 (1.001 – 1.003)** | **1.001 (1.001 – 1.002)** | **1.001 (1.001 – 1.002)** |

Using the random forests fitted for each ED, we observed that the Elixhauser score, age group, chief complaint, and insurance status were consistently at the top in terms of predictor importance (see Figure 1). The random forests had test accuracies of 68.9%, 72.1%, and 81.4% for EDs A, B, and C, respectively. As described above, we split the population according to the weighted Elixhauser score, age group, and insurance status.

**Figure 1.**
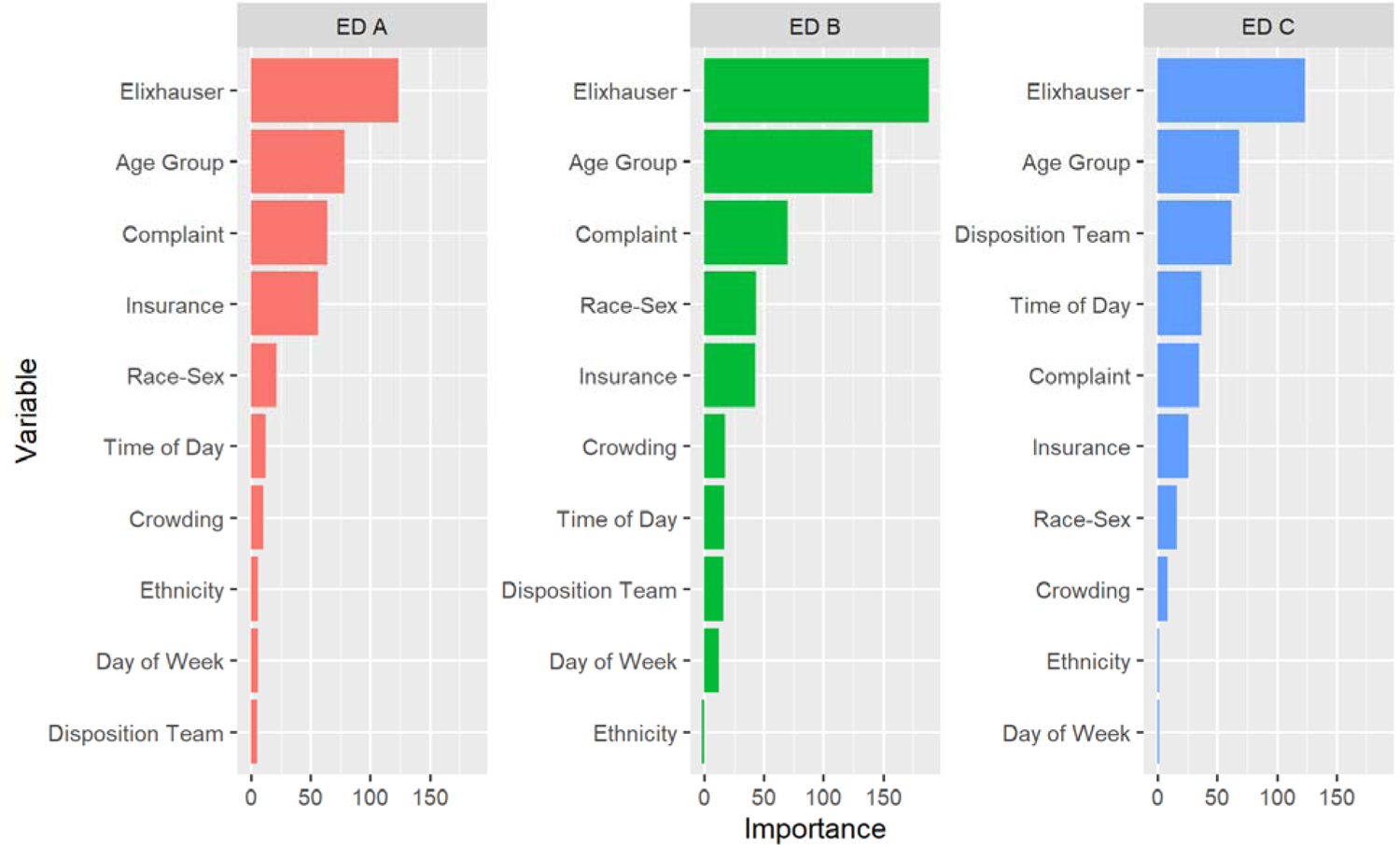
Variable importance plots based off mean decrease in accuracy for the three different random forests associated with the three EDs.

Figure 2 shows the odds ratio plots from the models formed by splitting the patient population according to weighted Elixhauser score. At all three EDs, African-American women were significantly less likely to be admitted than White men regardless of the Elixhauser score with odds ratios being as low as 0.45 for low Elixhauser category. African-American men and White women were also significantly less likely to be admitted if they had a low Elixhauser at all three hospitals. Within the middle Elixhauser group, the differences disappeared except for African-American women. In the high Elixhauser group, there was not a statistically significant difference between White women and White men at any of the EDs but both African-American men and women were less likely to be admitted compared to White men. These statistical insignificances observed in middle and high Elixhauser groups may be due to small sample sizes as more than 60, 80, and 70 percent of patient encounters in EDs A, B, and C, respectively, fell into the lower Elixhauser category, The next group of models explored how the admission odds changed between age groups. Figure 3 shows the associated odds ratio plots for the different age groups at each ED. Across all age groups, African-American women were less likely to be admitted compared to White men. When examining only White patients, the sex effect was reduced for older patients, culminating in White men and women with 65+ years of age having no significant differences in admission likelihood. (We do not suspect that this is due to a sample-size issue as the oldest age category has a similar sample size as the other categories in all EDs.) We also observed a diminished racial difference for younger males in EDs A and B, with White males and African-American males in the 18 through 40 group having no statistically significant difference in admission likelihood. Race-sex differences existed in the 41 through 64-year-old group across all three EDs.

**Figure 2.**
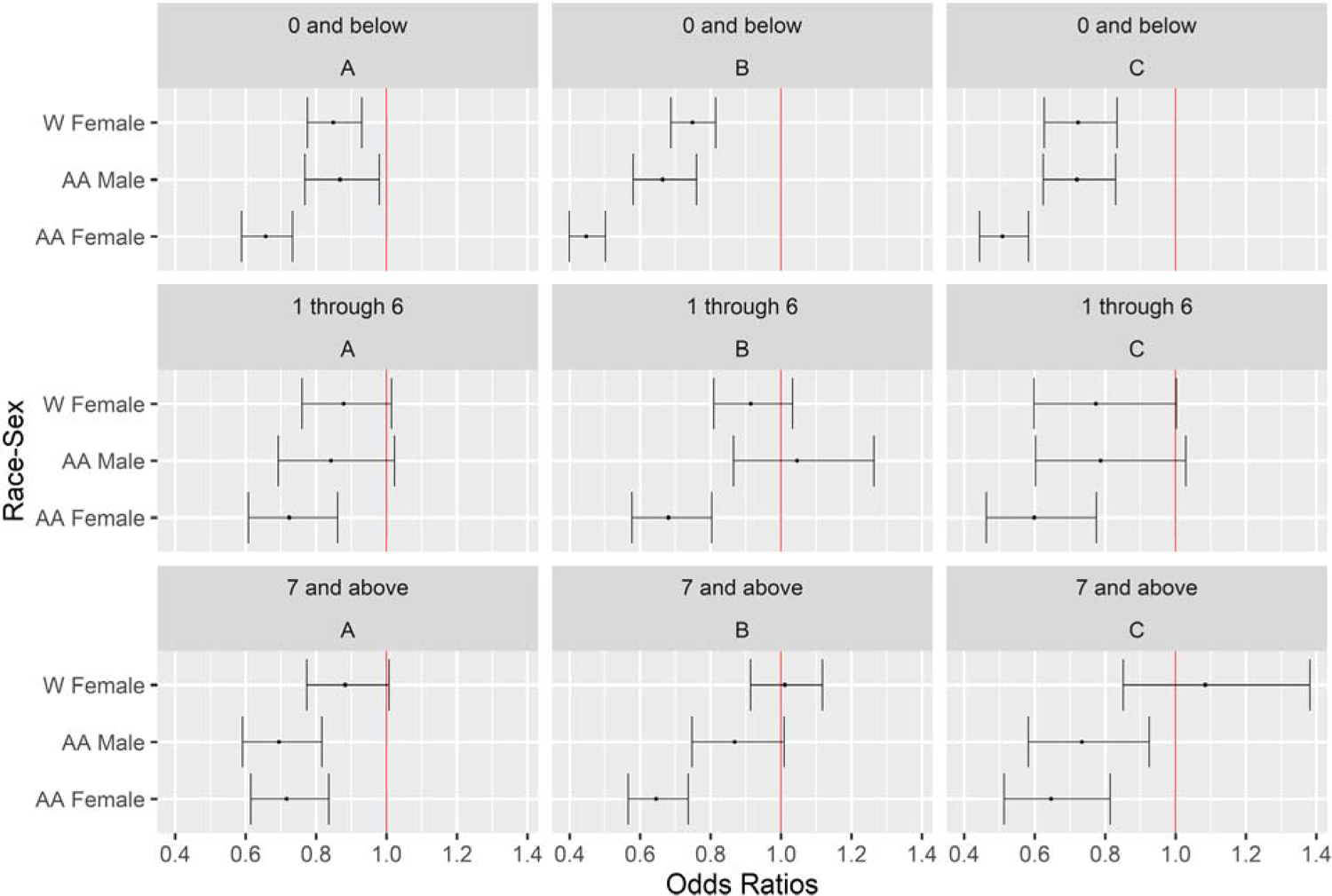
Odds ratio plots with 95% CIs for the Race-Sex variable for patients split by Elixhauser score at each ED, with the reference level being White men. Each row corresponds to a range of Elixhauser scores, and each column is an ED. On the vertical axis labels, AA means African-American, and W means White.

**Figure 3.**
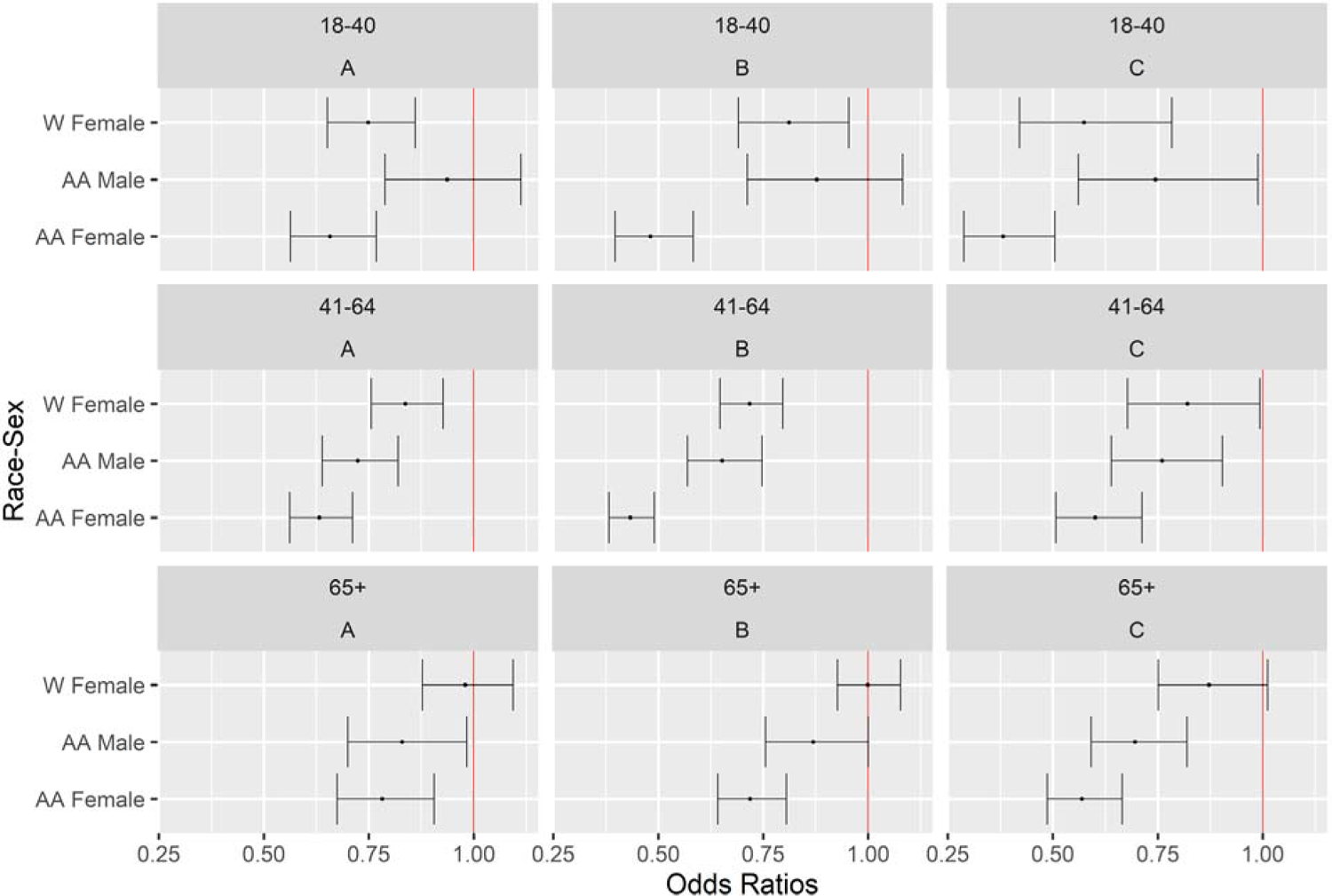
Odds ratio plots with 95% CIs for the Race-Sex variable for patients split by age group at each ED, with the reference level being White men. Each row corresponds to a different age group and each column is an ED.

The final group of models was formed using patients with the same insurance status. Figure 4 shows the odds ratios for each Race-Sex category for different insurance groups. Regardless of the insurance status, African-American women were less likely to be admitted than White men at all EDs except for Medicare patients at ED A. Considering all insurance groups, Medicare patients appear to be the least likely to experience race-sex differences in admission decisions. Within this group, no differences were observed for any Race-sex category except for African-American women at EDs B and C and for African-American men at ED C. (We do not suspect that this is due to a sample-size issue as the Medicare category is not a small category in comparison to other insurance categories in all EDs.) On the other hand, differences were prevalent in all other insurance groups except for White female and African-American male patients with Commercial/Other insurance at ED A, with Medicaid insurance at ED B, and who were in Self-Pay insurance category at ED C. (The indifferences for Medicaid and Self-pay patients in Figure 4 may be due to a statistical power issue as the confidence intervals for these cases are larger than others.)

**Figure 4.**
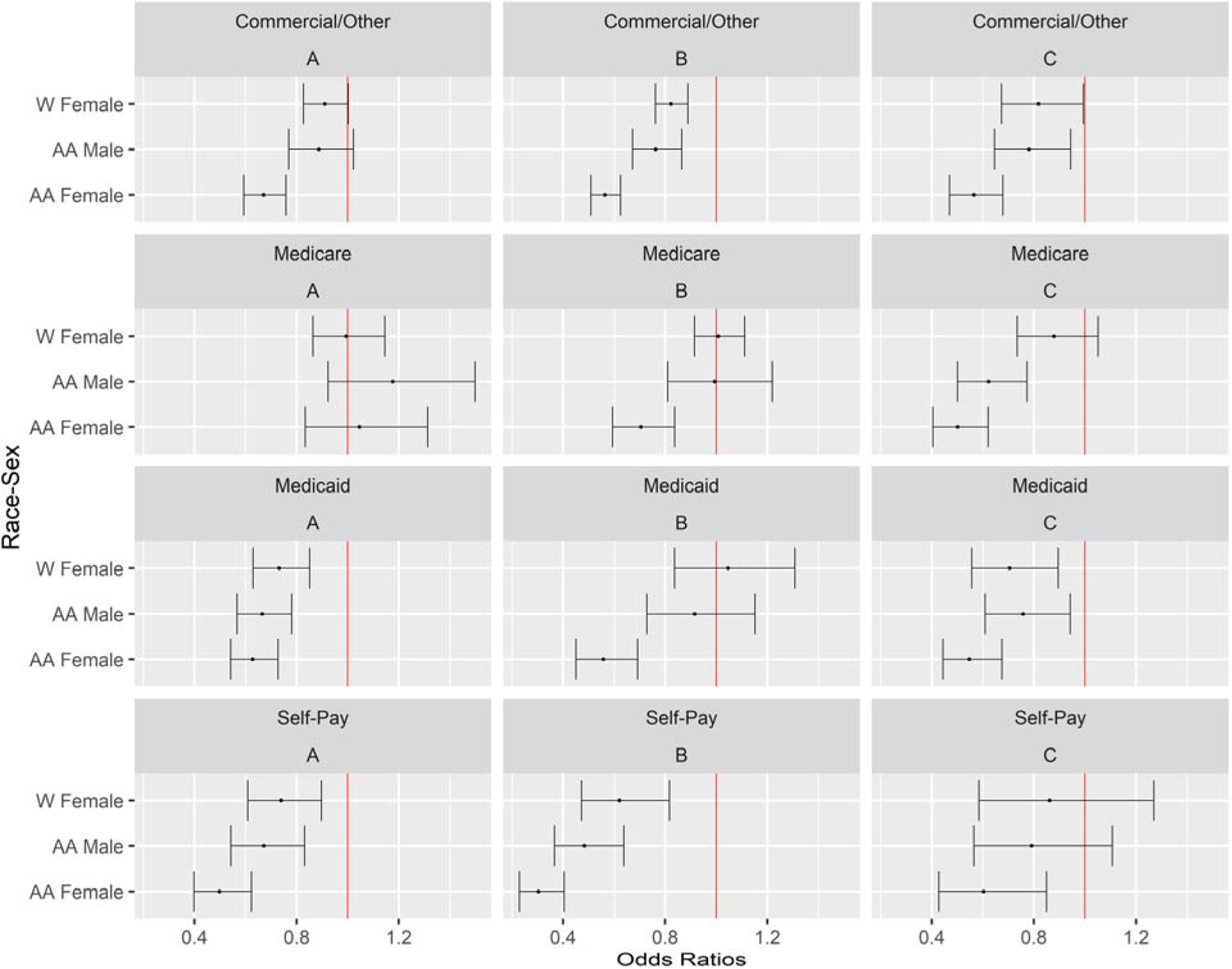
Odds ratio plots with 95% CIs for the Race-Sex variable for patients split by insurance status at each ED, with the reference level being White men. Each row corresponds to a different insurance group and each column is an ED.

## DISCUSSION

Our retrospective study of adult, intermediate-acuity ED patients found that African-American women were less likely to be admitted than White men at all three EDs regardless of subgroupings by comorbidity score, age, or insurance status (with the only exception being Medicare patients at ED A). Moreover, African-American women trended toward the lowest likelihood of admission compared to all other race-sex groups. In addition, we observed that African-American men also had lower likelihood of admissions compared to White men consistently across EDs within certain subpopulations. These findings raise further questions about the possibility of general presence of racial disparities within EDs as this pattern held regardless of whether the ED served predominantly White patients (EDs A and B) or African-American patients (ED C).

There have been several studies examining both patient and provider related factors that correlate with the disposition decision. For example, more senior physicians were found to have higher discharge rates^43^. On the patient side, women, African-Americans, and uninsured patients have been found to be less likely to be admitted from the ED compared to men, Whites, and privately insured patients, respectively, after accounting for age, acuity, comorbidities, and other factors^22–26^. Among heart failure patients, Whites have been found to be more likely to be admitted than African Americans^27^. Also, Maori patients in New Zealand were found to be less likely to be admitted than non-Maori patients who presented with similar triage acuities^28^. Our findings add further support to these preceding studies about the presence of disposition differences and how they affect different types of patients.

Our subgroup analyses of highly predictive factors identified by random forests suggest that differences were less prevalent for patients with the highest rates of admission: high Elixhauser, ages 65 years and older, and Medicare insurance. In particular, we note that the sex difference was less prevalent within high Elixhauser score patients as there was no significant difference between male and female White patients; we also observed overlapping confidence intervals for male and female African-American patients for this group. In addition, Medicare patients were the least likely group to experience race-sex differences in admission decisions.

According to our data, patients with low Elixhauser score, ages less than 65 years, and Self-pay were most prone to race-sex differences in ED disposition, although these results varied by ED. More specifically, the subpopulations that were most vulnerable to disparities in disposition decision in this study were (i) African-American women, (ii) African-American men who were 41+ years of age, and (iii) White women who were younger than 65 years of age, who had low comorbidity scores, or whose insurance status was Medicaid or Self-Pay.

Based on these observations of significant race-sex differences in ED admissions, our future research will investigate the causes and potential mechanisms of race-sex disparities in ED decision making. Although we were not able to distinguish between biological and social constructs in this study, observed differences in ED admission decisions by patient sex and race point to possible influences of implicit bias, systemic inequities, and other social factors. Further, we plan to explore potential solutions to these disparities tailored and targeted to patient subgroups (e.g., younger ages, fewer comorbidities, and uninsured) with the largest magnitude of race-sex differences.

## LIMITATIONS

This study had several limitations. Patient race and ethnicity were taken directly from what was recorded in the electronic health record system during clinical care instead of being prospectively collected. However, less than only 0.1% of all patient encounters had missing demographic information and furthermore, this information was collected by trained personnel. We did not have data on the sex and race of the clinicians making the disposition decisions, both of which could be important explanatory variables for a study on patient race-sex disparities.

We only considered White and African-American race categories as well as only male and female sex categories in this study, primarily due to insufficient sample sizes for other categories. We would need additional data from other sexes and races to further generalize our conclusions and how admission decisions are made with respect to other demographic groups.

Our models may not account for several unmeasured covariates that affect the admission decision-making process. For example, there are comorbidities that affect demographic groups differently such as end stage renal disease (ESRD) or sickle cell disease^44^. We do not explicitly account for these specific comorbidities that may impact the likelihood of admission.

We did not have data on patient outcomes following ED disposition and were not able to determine the most appropriate disposition decision for each patient. The goal of this study was to examine admission from the ED as a process measure to compare by patient sex and race. However, we acknowledge that discharge from the ED can be the more favorable decision for certain patients.

We did not conduct a power analysis for sample size calculations prior to the study. Therefore, some of the statistically insignificant differences observed may be due to insufficient sample sizes.

Our dataset did not include all information used by providers while making disposition decisions (such as entirety of patient chart), and the covariates used in this study could have been inconsistently recorded in the EHR. In particular, the existing diagnoses for certain patient groups could be undocumented in the EHR that would lead to underestimation of the Elixhauser comorbidity scores for that group. This compounded effect of biases at different stages of health care is not straightforward to study but is worthy of future research. Finally, this study considered three large EDs that are located within the Southeastern US. Therefore, our results should not be generalized to EDs with smaller patient populations and to EDs in other geographic locations with potentially different population demographics.

## CONCLUSIONS

We observed varying levels of differences in admission decisions for African-American men and women as well as White women when compared with White men within the adult ESI 3 patient populations from three large EDs in the Southeastern US. After stratifying on three patient characteristics highly predictive of admission in random forests, we identified consistent significant differences in disposition decisions by patient sex and race. Our findings indicated that certain differences disappeared or weakened for high Elixhauser score patients, Medicare patients, and older patients. In contrast, Medicaid and Self-Pay patients as well as those who had less severe comorbidities were more likely to experience lower likelihood of admissions. African-American women were consistently less likely to be admitted when compared to White men, even after splitting the data according to age, insurance status, or Elixhauser score. This finding in particular highlights urgent need for focused attention by policy makers and healthcare providers. Although results from this study should be only taken as indicative of differences in admission decisions for different demographic groups which does not directly translate to disparities, they still lead to important insights. Further research is needed to investigate whether the observed demographic differences in ED admissions are due to systemic or implicit biases.

### Statements and Declarations

PL, NTA, QC, CSE, BL, YL, AM, LM, MDP, and SZ report no conflict of interest. Although unrelated to the current work, CSE reports receiving personal consulting fees from Roche.

## Author Contributions

All authors conceived and designed the study; PL and QC managed the data; PL, NTA, QC, YL, MDP, and SZ analyzed numerical results; PL constructed the statistical models and drafted the manuscript; NTA, YL, MDP, and SZ provided statistical expertise; NTA, YL, MDP, and SZ acquired funding; NTA, YL, MDP, and SZ revised the manuscript; CSE, BL, AM, and LM provided clinical interpretations of the results and provided insights into the operational management of emergency department. All authors reviewed the manuscript and NTA takes responsibility for the paper as the corresponding author.

## Ethical Considerations

This study was reviewed by the IRB at the University of North Carolina at Chapel Hill and determined to be exempt from IRB review (UNC IRB **#:** 22-1714). As a retrospective study of existing data with limited identifiers (i.e., patient age and encounter date-times), informed consent was not required.

## Data Availability

Partial deidentifed patient-level data files and data dictionary are available upon request from 8/1/2024 by contacting Dr. Argon at to investigators with an IRB letter of approval and fully executed Carolina Data Warehouse for Health data use agreement with UNC Health.

## SUPPLEMENT

To ensure that there are no significant effects of multicollinearity in our logistic regression model for each ED, we calculated generalized variable inflation factors (GVIF), see, e.g., Fox and Monette (1992). For variables with one degree of freedom, the GVIF is equivalent to the variable inflation factor (VIF) whereas for variables with two or more degrees of freedom, calculating 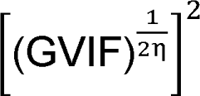 allows us to use the same VIF thresholds as for continuous variables (i.e., those with one degree of freedom). Table S1 shows that the highest value for this calculation is approximately 1.295, which is below the recommended threshold of 2. Therefore, we conclude that multicollinearity is not a concern for either model.

**Table S1.** 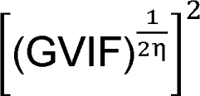 values for logistic regression models, where η is the degrees of freedom associated with each predictor. These are directly comparable to VIF thresholds used for continuous variables.

| **Predictor** | **ED A** | **ED B** | **ED C** |
| --- | --- | --- | --- |
| Age Group | 1.237 | 1.295 | 1.285 |
| Race-Sex | 1.046 | 1.064 | 1.051 |
| Ethnicity | 1.005 | 1.003 | 1.003 |
| Complaint Category | 1.028 | 1.011 | 1.012 |
| Insurance | 1.155 | 1.184 | 1.175 |
| Elixhauser | 1.124 | 1.157 | 1.111 |
| Disposition Care Area | 1.111 | 1.030 | 1.062 |
| Time of Day | 1.056 | 1.045 | 1.024 |
| Day of Week | 1.014 | 1.003 | 1.007 |
| Crowding | 1.275 | 1.180 | 1.059 |

**Table S2.**
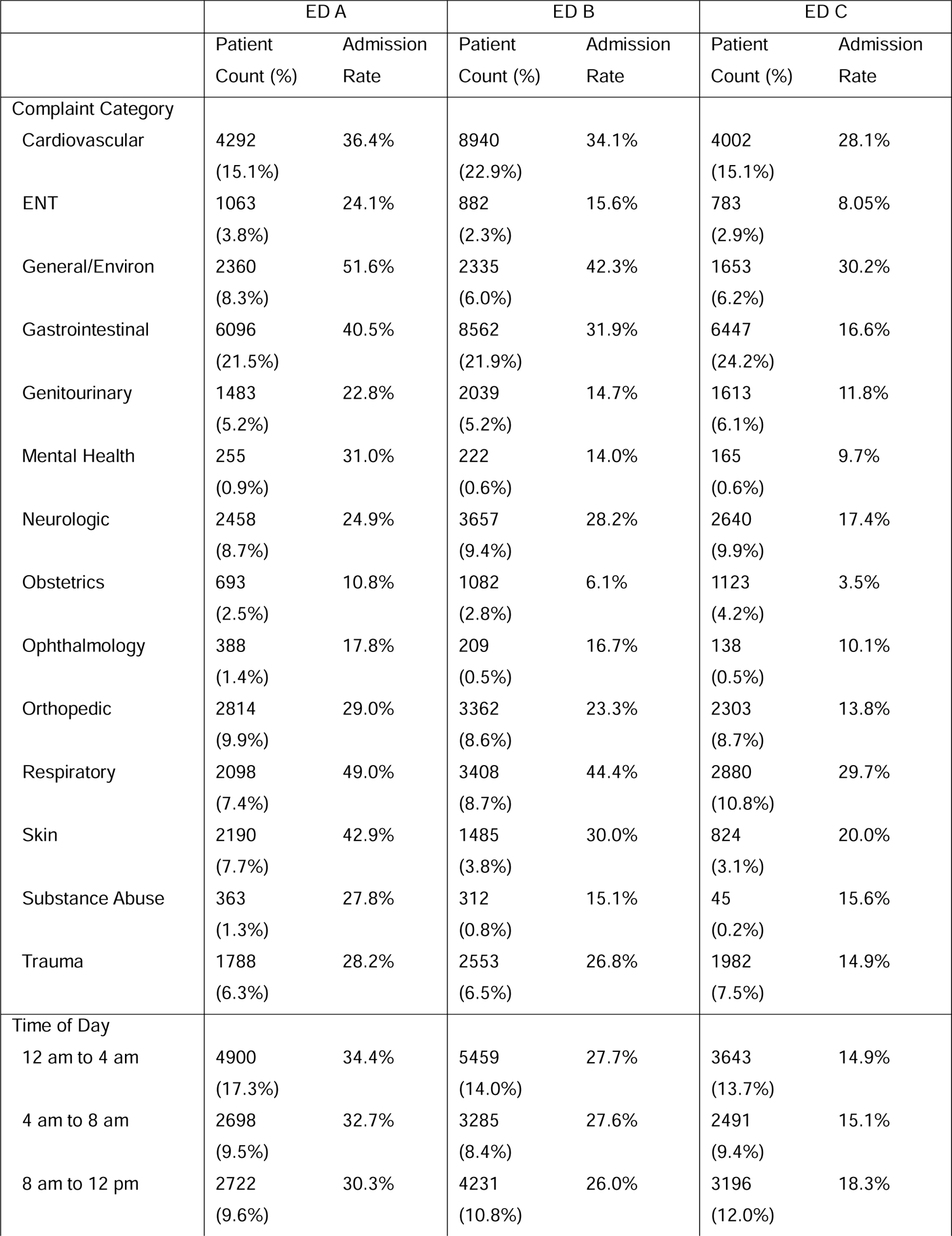
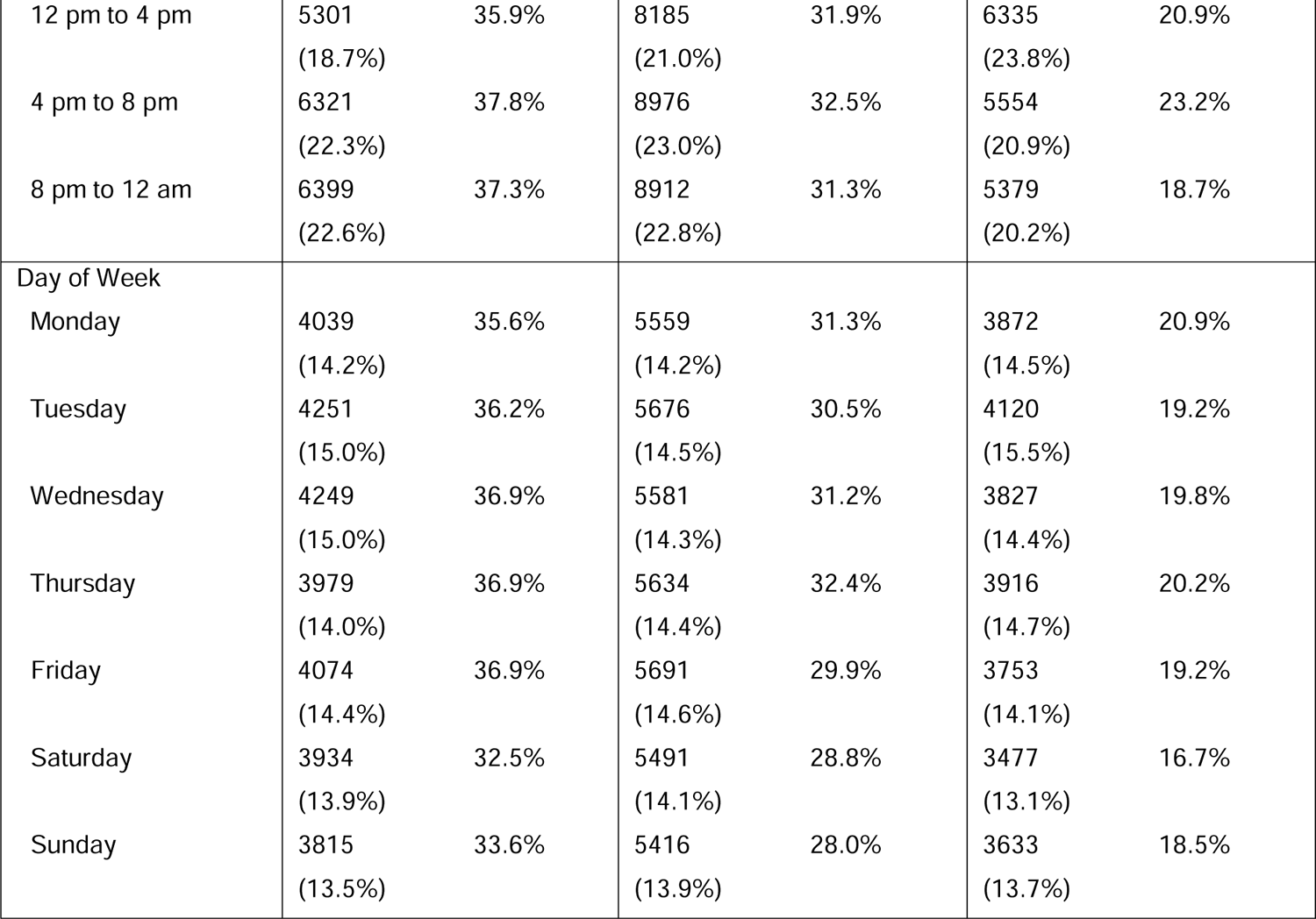
Additional breakdown of study patient characteristics with the same columns as Table 1.

|  | ED A |  | ED B |  | ED C |  |
| --- | --- | --- | --- | --- | --- | --- |
|  | Patient<br>Count (%) | Admission<br>Rate | Patient<br>Count (%) | Admission<br>Rate | Patient<br>Count (%) | Admission<br>Rate |
| Complaint Category |  |  |  |  |  |  |
| Cardiovascular | 4292<br>(15.1%) | 36.4% | 8940<br>(22.9%) | 34.1% | 4002<br>(15.1%) | 28.1% |
| ENT | 1063<br>(3.8%) | 24.1% | 882<br>(2.3%) | 15.6% | 783<br>(2.9%) | 8.05% |
| General/Environ | 2360<br>(8.3%) | 51.6% | 2335<br>(6.0%) | 42.3% | 1653<br>(6.2%) | 30.2% |
| Gastrointestinal | 6096<br>(21.5%) | 40.5% | 8562<br>(21.9%) | 31.9% | 6447<br>(24.2%) | 16.6% |
| Genitourinary | 1483<br>(5.2%) | 22.8% | 2039<br>(5.2%) | 14.7% | 1613<br>(6.1%) | 11.8% |
| Mental Health | 255<br>(0.9%) | 31.0% | 222<br>(0.6%) | 14.0% | 165<br>(0.6%) | 9.7% |
| Neurologic | 2458<br>(8.7%) | 24.9% | 3657<br>(9.4%) | 28.2% | 2640<br>(9.9%) | 17.4% |
| Obstetrics | 693<br>(2.5%) | 10.8% | 1082<br>(2.8%) | 6.1% | 1123<br>(4.2%) | 3.5% |
| Ophthalmology | 388<br>(1.4%) | 17.8% | 209<br>(0.5%) | 16.7% | 138<br>(0.5%) | 10.1% |
| Orthopedic | 2814<br>(9.9%) | 29.0% | 3362<br>(8.6%) | 23.3% | 2303<br>(8.7%) | 13.8% |
| Respiratory | 2098<br>(7.4%) | 49.0% | 3408<br>(8.7%) | 44.4% | 2880<br>(10.8%) | 29.7% |
| Skin | 2190<br>(7.7%) | 42.9% | 1485<br>(3.8%) | 30.0% | 824<br>(3.1%) | 20.0% |
| Substance Abuse | 363<br>(1.3%) | 27.8% | 312<br>(0.8%) | 15.1% | 45<br>(0.2%) | 15.6% |
| Trauma | 1788<br>(6.3%) | 28.2% | 2553<br>(6.5%) | 26.8% | 1982<br>(7.5%) | 14.9% |
| Time of Day |  |  |  |  |  |  |
| 12 am to 4 am | 4900<br>(17.3%) | 34.4% | 5459<br>(14.0%) | 27.7% | 3643<br>(13.7%) | 14.9% |
| 4 am to 8 am | 2698<br>(9.5%) | 32.7% | 3285<br>(8.4%) | 27.6% | 2491<br>(9.4%) | 15.1% |
| 8 am to 12 pm | 2722<br>(9.6%) | 30.3% | 4231<br>(10.8%) | 26.0% | 3196<br>(12.0%) | 18.3% |
| 12 pm to 4 pm | 5301<br>(18.7%) | 35.9% | 8185<br>(21.0%) | 31.9% | 6335<br>(23.8%) | 20.9% |
| 4 pm to 8 pm | 6321<br>(22.3%) | 37.8% | 8976<br>(23.0%) | 32.5% | 5554<br>(20.9%) | 23.2% |
| 8 pm to 12 am | 6399<br>(22.6%) | 37.3% | 8912<br>(22.8%) | 31.3% | 5379<br>(20.2%) | 18.7% |
| Day of Week |  |  |  |  |  |  |
| Monday | 4039<br>(14.2%) | 35.6% | 5559<br>(14.2%) | 31.3% | 3872<br>(14.5%) | 20.9% |
| Tuesday | 4251<br>(15.0%) | 36.2% | 5676<br>(14.5%) | 30.5% | 4120<br>(15.5%) | 19.2% |
| Wednesday | 4249<br>(15.0%) | 36.9% | 5581<br>(14.3%) | 31.2% | 3827<br>(14.4%) | 19.8% |
| Thursday | 3979<br>(14.0%) | 36.9% | 5634<br>(14.4%) | 32.4% | 3916<br>(14.7%) | 20.2% |
| Friday | 4074<br>(14.4%) | 36.9% | 5691<br>(14.6%) | 29.9% | 3753<br>(14.1%) | 19.2% |
| Saturday | 3934<br>(13.9%) | 32.5% | 5491<br>(14.1%) | 28.8% | 3477<br>(13.1%) | 16.7% |
| Sunday | 3815<br>(13.5%) | 33.6% | 5416<br>(13.9%) | 28.0% | 3633<br>(13.7%) | 18.5% |

